# Effect of high-intensity interval training on body composition and glucose control in type 2 diabetes: a systematic review and meta-analysis

**DOI:** 10.1101/2023.01.27.23285090

**Authors:** Yidan Cai, Zhuan Long, Zhicheng Guo

## Abstract

**Background:** Diabetes mellitus(DM) has become the third chronic noncommunicable disease worldwide, and is one of the most common chronic diseases in almost all countries. Type 2 diabetes(T2DM) is the most common group of DM, accounting for more than 90% of the DM population.

**Objectives:** This systematic review is conducted to compare the impact of HIIT and MICT on body composition and glucose control in T2DM, and to determine the suitable intervention for HIIT and the more effective forms of HIIT on T2DM.

**Methods:** Seven databases were searched from their inception to 21 November 2022 for randomized controlled trial(RCT) with HIIT and MICT intervention. The effect size was completed by using standardized mean difference (SMD) and standard deviation. Body mass(BM), body mass index (BMI), percent fat mass (PFM), fat mass (FM), fat-free mass (FFM), VO_2peak_, HbA1c, fasting plasma glucose(FPG), and fasting plasma insulin(FPI) were included in the meta-analysis as outcomes.

**Results:** 15 RCTs with 371 T2DM were conducted in accordance with our inclusion criteria. The results of the meta-analysis revealed that compared to MICT, HIIT had significant effects on VO_2peak_(SMD=0.4, 95%CI: 0.08 to 0.73, p=0.02) and HbA1c(SMD=-0.24, 95%CI: -0.48 to -0.01, p=0.04), while there were no significant differences in body composition, FPG, and FPI.

**Conclusion:** HIIT provides similar or more benefits on body composition, cardiorespiratory fitness(CRF), and glucose control relative to MICT, which might be influenced by duration, frequency, and HIIT interval. For people with T2DM, HIIT can achieve more improvement in CRF and glucose control than MICT and appear to be more time-saving.

## 1. Introduction

Diabetes mellitus(DM) is a metabolic disorder characterized by elevated blood glucose and uncontrollable disease. Polyuria, polydipsia, polyphagia and weight loss are common clinical manifestations of DM. Studies have found that, except for cardiovascular and cerebrovascular diseases and tumors, DM has become the third chronic noncommunicable disease worldwide, and is one of the most common chronic diseases in almost all countries^[1]^. As economic development and urbanization leading to lifestyle changes characterized by decreased physical activity and increased obesity, the number of patients with diabetes is increasing year by year^[2]^.

T2DM is the most common group of DM, accounting for more than 90% of the DM population^[3]^. Due to uncontrollable blood glucose and high blood lipid, T2DM patients can induce cardiovascular disease and lead to cardiovascular deterioration, which seriously threatens the quality of life of T2DM patients^[4]^. Moreover, the growing economy of treating T2DM has caused burdens on individuals, families and society, so effective and economic exercise intervention is particularly important^[5]^. Exercise therapy, as one of the new “five carriages” for the treatment of T2DM, has already attracted wide attention. Adherence to regular aerobic exercise is considered an important tool for the prevention and management of T2DM and its complications, improving the insulin sensitivity of human body, and promote T2DM patients to better control blood sugar and blood fat, improve their cardiopulmonary endurance, and improve / slow down the emergence and development of long-term complications^[6]^.

Moderate-intensity continuous aerobic training (MICT) is a routine aerobic exercise method in patients with T2DM. However current exercise guidelines require T2DM to perform MICT for at least 150min per week, which is hard to adhere to^[7]^. High-intensity interval training (HIIT) usually performs multiple short-time, high-intensity exercises, interspersed with intermittent, short-time, low-intensity or complete rest, periodic cycle of exercise. HIIT was found to be effective in improving blood glucose and body composition in patients with T2DM^[8]^. HIIT can significantly reduce the duration of exercise by improving the intensity of exercise, that is, reduce the amount of exercise, and can effectively promote lactate clearance, relieve fatigue and discomfort of patients, and reduce the occurrence of adverse events of patients^[9]^. Robinson^[10]^ et al. performed HIIT in T2DM patients with 1min85% -90% peak heart rate (HRpeak) intensity and intermittent 1min of low intensity exercise, and after 2 weeks of protocol intervention, the results found that both HIIT and MICT could improve glycemic fructosamine concentration and effectively control blood glucose increase. Ramos^[11]^ et al found that only HIIT reduced insulin levels in non-T2DM patients with metabolic syndrome, but not in fasting plasma proinsulin levels in T2DM patients before intervention. A recent systemtic review also found that there were no significant differences in the HIIT group in total cholesterol (TC) and HDL cholesterol (HDL-C), fat mass(FM), waist circumference, and hemoglobin A1c (HbA1c)^[12]^. In summary, It is still controversial whether HIIT intervention can improve body composition, insulin sensitivity, and HbA1c in T2DM patients compared with MICT.

Over the last decade, several studies took glucose control as the core subjects in exploring the effect of HIIT intervention, however, ignored the relation between body composition and glucose control. And as core measures of exercise effect, the improvement of body composition and cardiorespiratory fitness(CRF) are inseparable and need to be explored with glucose control together in people with T2DM. Despite clear evidence for the benefit following HIIT compared with MICT on body composition and CRF, there is no consensus on whether HIIT is similar or superior to MICT on body composition and glucose in people with T2DM. This systematic review was conducted to compare the impact of HIIT and MICT on body composition and glucose control in T2DM, and to determine the suitable intervention for HIIT and the more effective forms of HIIT on T2DM.

## Method

This systematic review was registered in the International Prospective Register of Systematic Reviews(PROSPERO). The registration number was CRD42021285158.

### Literature search strategy

The review was conducted following the guidelines of the PRISMA-P statement. A throughout search of the electronic literature was carried out up to 21 December 2022, including Pubmed, Embace, the Cochrane Library, Web of Science, CNKI, and Wanfang. The search criteria were developed through some similar systematic reviews’, ‘high-intensity interval training, ‘type 2 diabetes, and so on were chosen as the key phrases. Searching results were imported into a reference manager(Endnote 20). To make sure more relevant studies were included in the review, we also examined the references list of the eligible studies. The papers were appraised by two researchers independently. By thorough screening, the papers that fitted our criteria remained. Any disputes were solved by a third researcher through conversation.

### Inclusion and Exclusion Criteria

The following PICOS criteria were used on the screen:

#### Participants

Patients who were diagnosed with type 2 diabetes or pre-diabetes were included in this study, and the mean age of the participants in eligible studies was not limited. The subjects were limited to human, animal-based and age-incompatible subjects were excluded. There was no restriction on participants with medical comorbidities in the review.

#### Intervention

The intervention of the participants was HIIT only. The intensity of HIIT was measured between 80-100% HR_max_ or VO_2peak_, or at maximum effort, or a rating of perceived exertion(RPE) greater than 15. The duration of HIIT was a minimum of 2 weeks, and the passive recovery or low-intensity exercise in HIIT was between the 1min and 4min. The review did not limit the form of HIIT, but HIIT combined with other interventions(e.g., resistance training) was excluded.

#### Comparison

The included studies comprised a comparator group that undertook MICT. The training programs of MICT were at intensity 40 to 80% HR_max_ or VO2_peak_, or an RPE between 12-15, and the duration was over 15 min. There was also no restriction on the form of MICT.

#### Outcomes

The primary data related to body composition(BM, body mass index (BMI), waist circumference (WC), fat mass (FM), fat-free mass (FFM), percent fat mass (PFM)), Glycosylated Hemoglobin, Type A1C(HbA_1c_), fasting plasma glucose(FPG), fasting plasma insulin(FPI), and cardiorespiratory fitness(VO_2peak_) were included. The outcomes were all directly reported in the studies, the recalculated values were excluded.

#### Study

Research involving randomized controlled trials (RCT) written in English and Chinese. Observational studies, reviews, and studies and abstracts without adequate data were excluded.

### Data extraction

Two researchers extracted data independently, and the extracted results were checked by another two researchers. The basic characteristics of studies including age, sex ratio, the health status of participants, form of intervention, participants population, intervention characteristics(intensity, duration, and frequency of HIIT and MICT), and dropouts(both HIIT and MICT) were extracted. The mean ± standard deviation values, mean difference(MD), and 95% confidence intervals(95%CI) of pre-intervention, post-intervention, and changes between pre and pro-intervention(if reported) were extracted. When the outcomes reported by studies were insufficient or hard to extract, we would contact the corresponding authors for the data needed in the meta-analysis.

### Study quality assessment

The appraisal of the included studies was conducted by two reviewers. Considering HIIT and MICT are a form of physiotherapy, a Physiotherapy Evidence Database (PEDro) scale was used to assess the quality. The encompassing external validity (1 item), internal validity (8 items), and statistical reporting (2 items) of the eligible studies were checked to assess the quality. All items are rated yes or no according to whether the criterion is satisfied in the study.

### Statistical analysis

In this review, all analyses were carried out using the Revman 5.4 and Stata 15. The mean and standard deviation of the changes between pre-intervention and post-intervention in the HIIT and MICT groups was used to compare the between-group differences. If the mean and standard deviation of the changes were not reported directly, we would calculate them through pre-intervention and post-intervention values. Considering some experimental endpoints were highly variable, we adopted a random-effects model for all outcomes. The standardized mean difference (SMD) with 95% CI would be used to complete the effect size(ES). The heterogeneity among studies was quantified using Cochran’s Q test and the inconsistency I^2^ test. When I^2^ was 0 to 50%, the heterogeneity was considered to be acceptable. The funnel plots and Egger’s test was adopted to assess the publication bias.

To test the sensitivity, we carried out several subgroup analyses to find out whether the individual characteristics or intervention characteristics of each eligible study can influence the final result. Duration(<3 and ≥3 months), frequency(≤3 and >3 times/week), and interval protocol(<3 and ≥3 mins) were examined as subgroups.

## Results

### This Included studies

As shown in Figure 1, the search strategy retrieved 920 studies from electronic databases and 104 studies from references and other sources. After removing the duplicates, 1024 studies were evaluated via title and abstract, and 269 studies remained to be full-text screened. After diligently reviewing, 254 studies were removed for not meeting the inclusion criteria. Finally, 15 studies were evaluated as eligible and included in this analysis.

**Figure 1.**
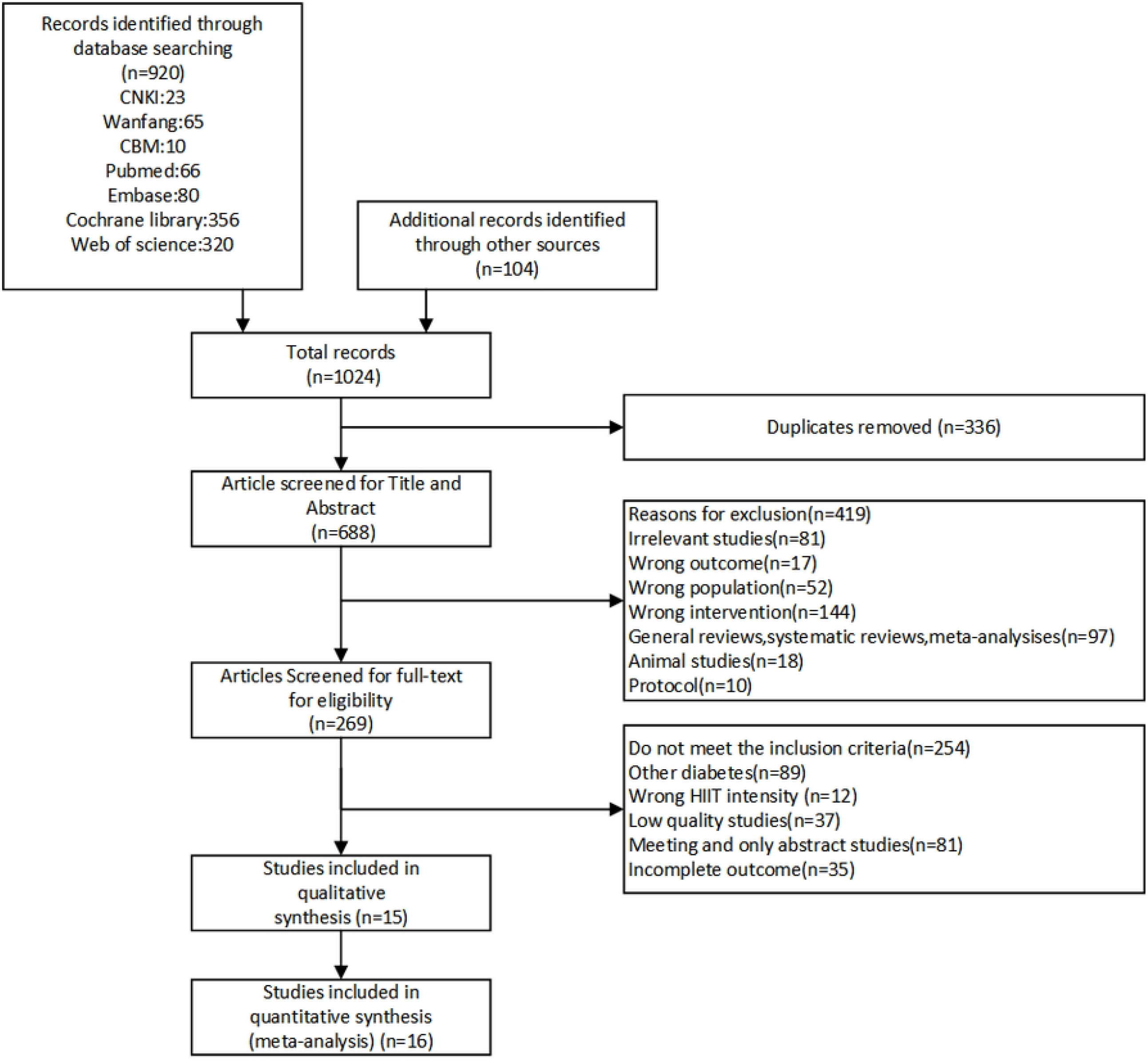
Literature search and study selection process.

#### Participant and Intervention characteristics

A total of 371 participants in 15 studies[13],[14],[15], [16],[17],[18],[19],[20],[21],[22],[23],[24],[25],[26],[27]were included in our meta-analysis. The primary characteristics of the participant and intervention are summarized in Table 1. 188 participants were allocated to the HIIT group, and 183 participants were allocated to the MICT group. Of all interventions, Most exercise forms were cycling, 3 studies used walking, 1 study used both cycling and running.

**Table 1.**
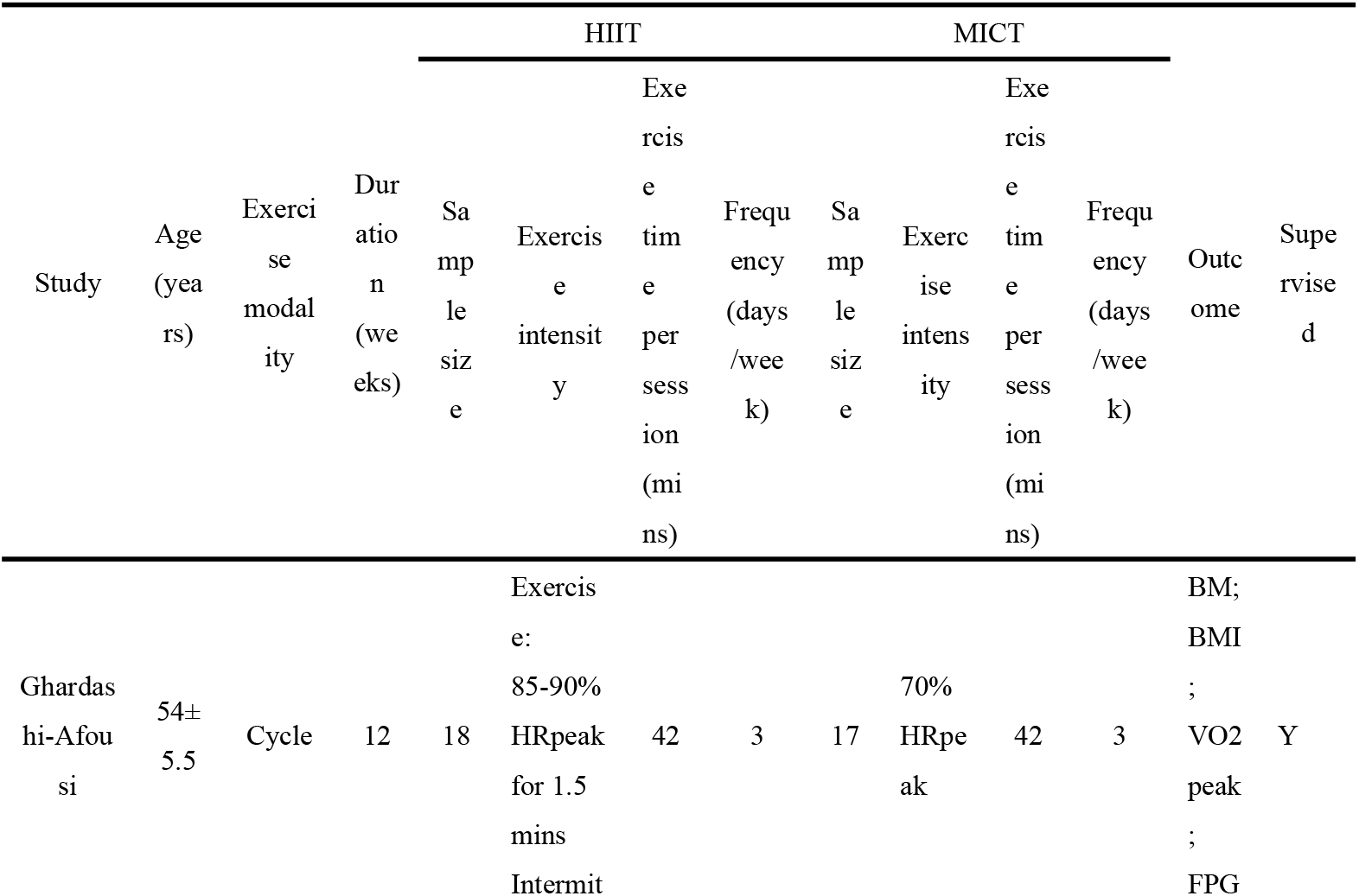
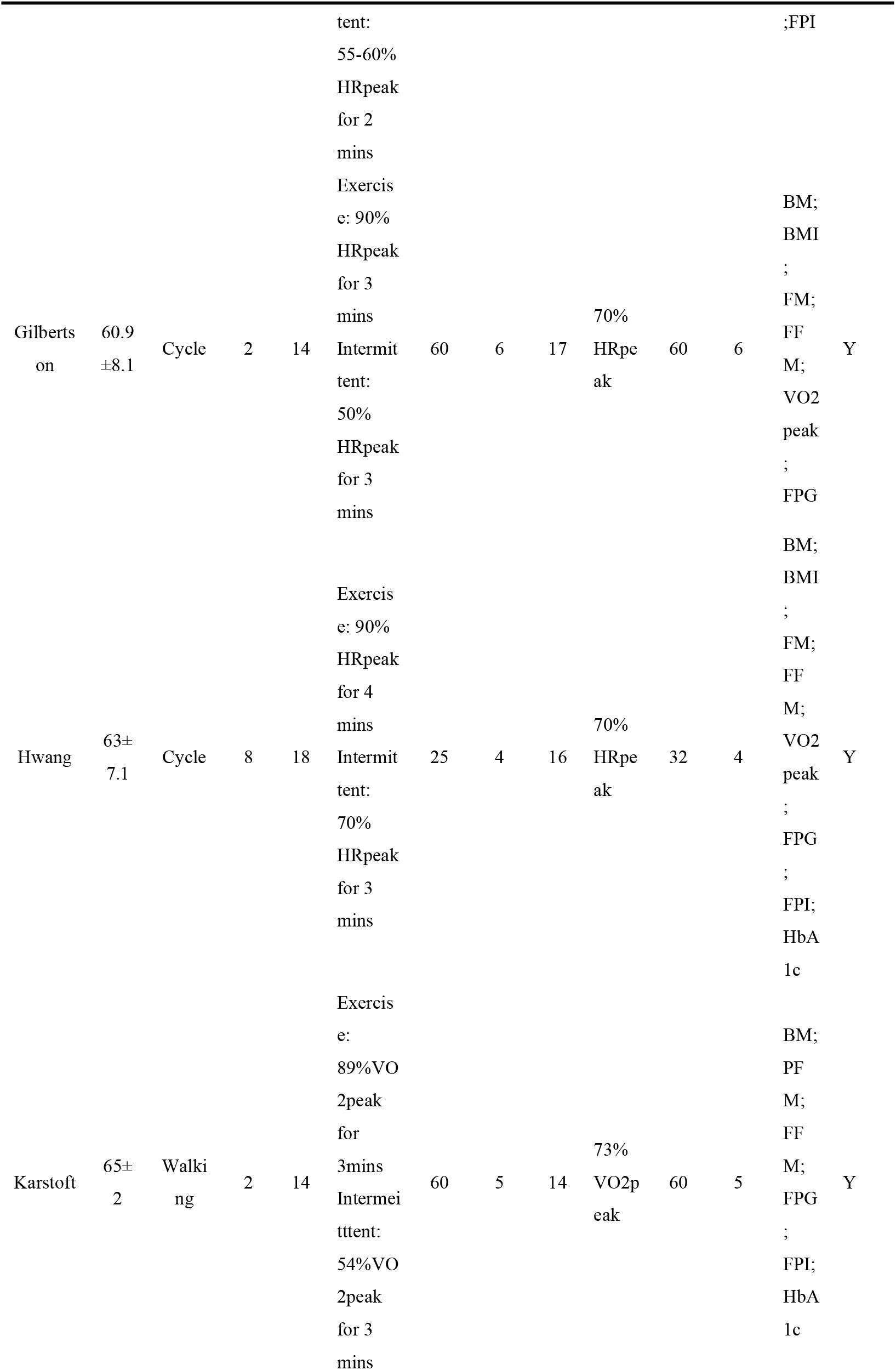
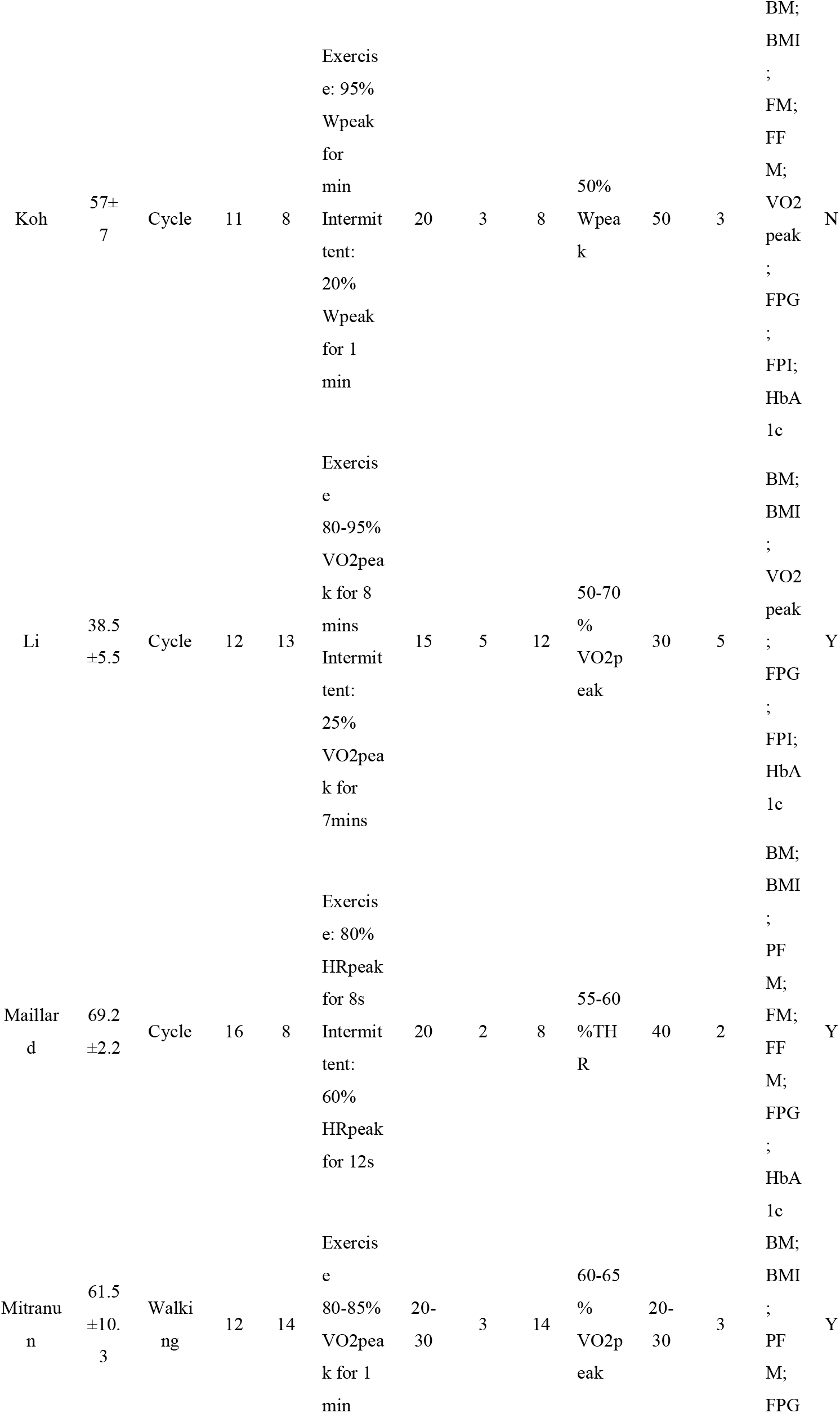
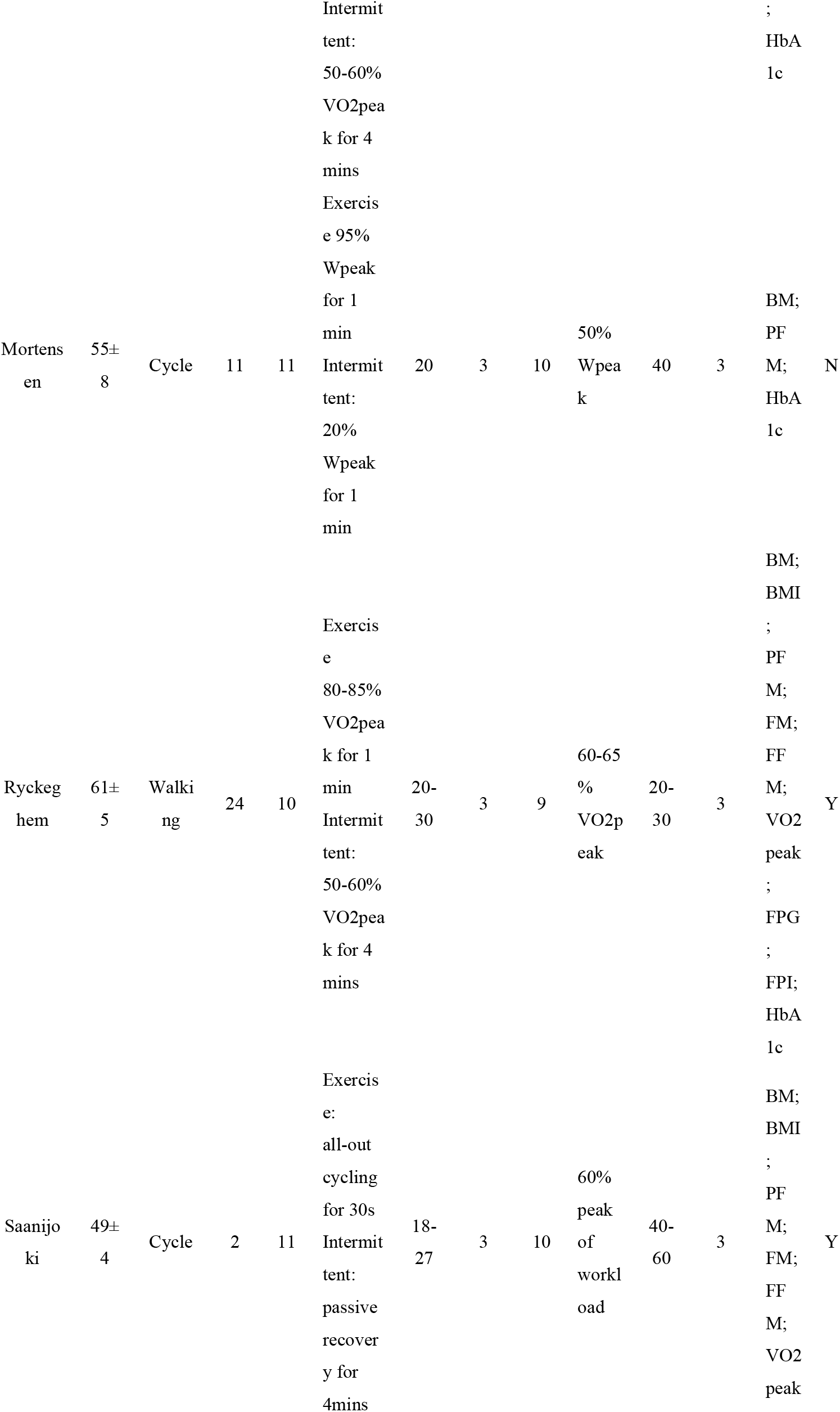
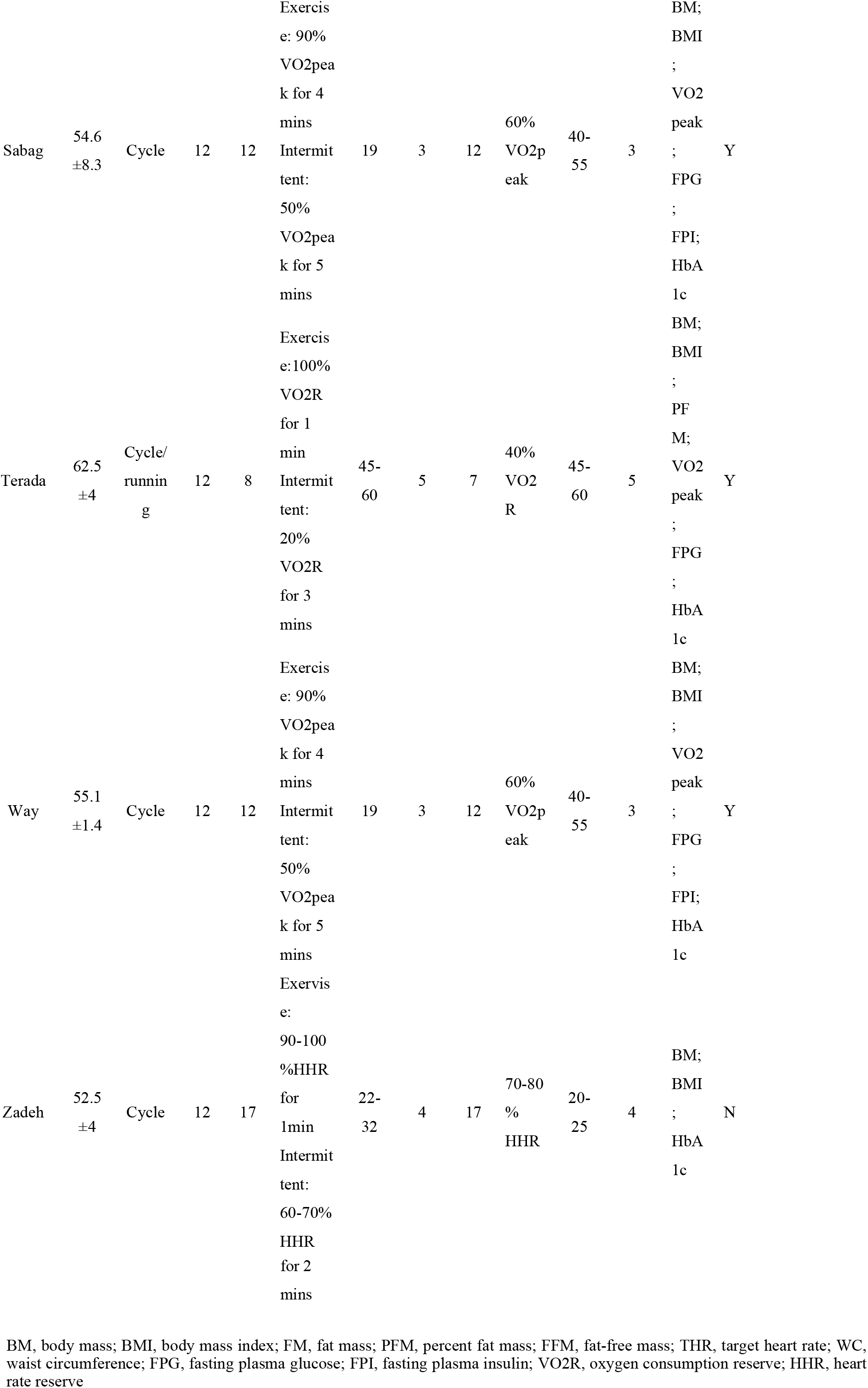
Characteristics of included studies.

The duration of 7 studies was 12 weeks, only 2 studies adopted > 3 weeks intervention. Most interventions used HRmax, Wpeak or VO2peak to measure the intensity of exercise, 1 study used THR(target heart rate), 1 study VO2R and 1 study HHR(heart rate reserve). Only 1 HIIT intervention used passive recovery, the remaining used active recovery. The exercise time ranged from 15 to 60 mins for HIIT and 30 to 60 mins for MICT. 8 studies instructed participants to exercise 3 times/week, 5 studies instructed > 3 times/week, and 2 studies instructed 2 time/week. 7 studies had > 15% dropout rate. 3 training interventions were not supervised.

### Quality assessment

As Table 2 presented, the quality of included studies was moderate(mean±SD = 5.07±1.06). All studies randomly allocated participants. 10 studies used concealed allocation. Only 1 study did not report and compare the baseline data of the participants. Because of the characteristic of exercise intervention, no studies blinded subjects or therapists. For assessors, most studies did not report the blind method or blind assessors, and only 2 studies blinded the assessors. 7 studies lost more than 15% of participants to follow-up. 8 studies reported adequate information on intention. All studies reported between-group statistics. 10 studies did not reported point measures.

**Table 2.**
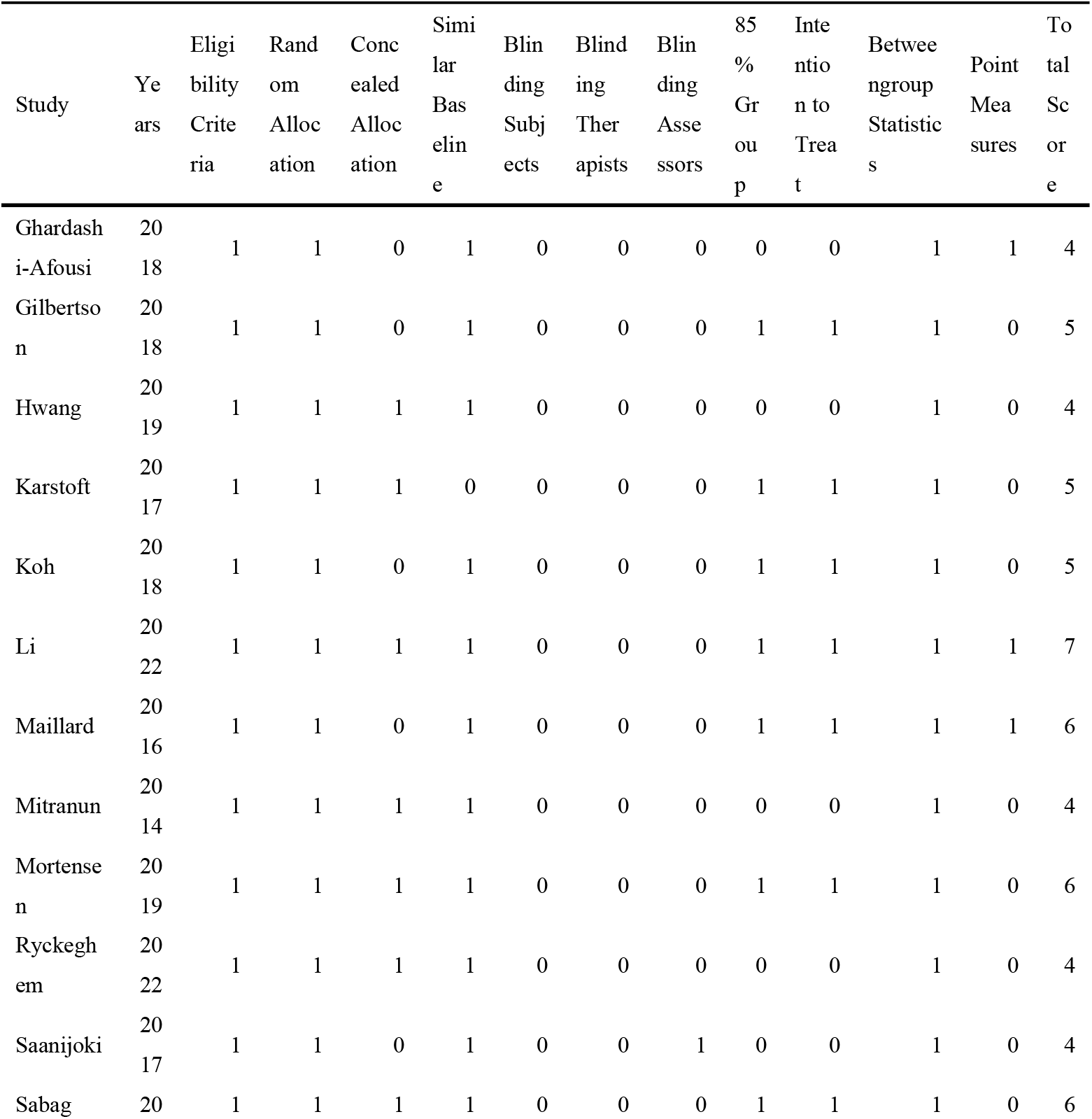
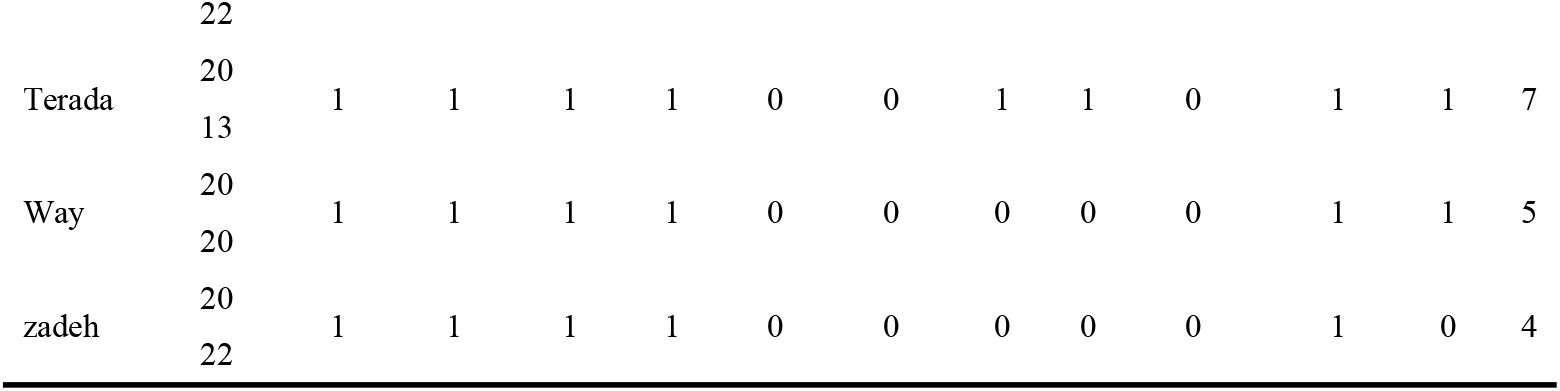
Quality assessment of study quality.

### Meta-analysis

As summarized in figure 2, All studies reported pre- and post-intervention in BM, 13 studies reported BMI, 8 studies reported PFM, 5 studies reported FM, 7 studies reported FFM, 12 studies reported HbA1c, 12 studies reported FPG, and 9 studies reported FPI. For cardiorespiratory fitness 11 studies reported VO2peak. The analyses were completed by using the above data. The analyses of HIIT vs MICT on T2DM were presented in Figure 2. There were significant differences between HIIT and MICT in HbA1c (SMD=-0.24, 95%CI: -0.48 to -0.01, p=0.04), and VO2peak (SMD=0.4, 95%CI: 0.08 to 0.73, p=0.02). No statistical differences were found in BM (SMD=0.06, 95%CI: -0.21 to 0.33, p=0.65), BMI (SMD=0.05, 95%CI: -0.23 to 0.33, p=0.74), PFM (SMD=0.02, 95%CI: -0.27 to 0.32, p=0.87), FM (SMD=0.04, 95%CI: -0.33 to -0.4, p=0.85), FFM (SMD=-0.03, 95%CI: -0.33 to 0.28, p=0.86), FPG (SMD=0.05, 95%CI: -0.18 to 0.28, p=0.65), and FPI (SMD=-0.18, 95%CI: -0.47 to 0.11, p=0.23).

### Subgroup analysis

As shown in the Table 3, according to the different characteristics of included studies, studies were divided into 6 subgroups. As the result showed, no matter duration, frequency, and HIIT interval, there was no statistical difference on BM, BMI, PFM, FM, FFM, FPG, and FPI. On HbA1c, a significant effect of duration(<3 months vs ≥3 months) was found in the subgroup analysis. Subgroup analysis of VO2peak identified a significant effect of frequency(≤3 times/week vs >3 times/week), and HIIT interval(1-3 mins vs ≥ 3 mins) between HIIT and MICT.

**Table 3.** Summary of HIIT vs MICT subgroup meta-analysis.

| Outcome | Studies(n) | SMD | (95%CI) | p | Heterogeneity |  |
| --- | --- | --- | --- | --- | --- | --- |
|  |  |  |  |  | I2(%) | p |
| BM |  |  |  |  |  |  |
| Duration:< 3 months | 6 | 0.15 | -0.32, 0.63 | 0.52 | 65% | 0.003 |
| Duration:≥ 3 months | 9 | -0.03 | -0.35, 0.29 | 0.87 | 0% | 1 |
| Frequency:≤ 3 times/week | 9 | 0.03 | -0.25, 0.3 | 0.83 | 0% | 1 |
| Frequency:> 3 times/week | 6 | 0.2 | -0.5, 0.89 | 0.58 | 78% | 0.0003 |
| HIIT interval: 1-3mins | 9 | -0.09 | -0.37, 0.18 | 0.51 | 0% | 0.74 |
| HIIT interval: ≥3mins | 6 | 0.3 | -0.28, 0.88 | 0.31 | 70% | 0.005 |
| BMI |  |  |  |  |  |  |
| Duration:< 3 months | 4 | -0.02 | -0.4, 0.37 | 0.94 | 0 | 1 |
| Duration:≥ 3 months | 9 | 0.1 | -0.32, 0.51 | 0.65 | 56% | 0.02 |
| Frequency:≤ 3 times/week | 8 | -0.02 | -0.31, 0.27 | 0.9 | 0% | 0.96 |
| Frequency:> 3 times/week | 5 | 0.21 | -0.5, 0.91 | 0.57 | 75% | 0.003 |
| HIIT interval: 1-3mins | 8 | -0.16 | -0.45, 0.13 | 0.29 | 0% | 0.83 |
| HIIT interval: ≥3mins | 5 | 0.35 | -0.23, 0.94 | 0.23 | 64% | 0.02 |
| FAT% |  |  |  |  |  |  |
| Duration:< 3 months | 4 | 0.03 | -0.35, 0.42 | 0.87 | 0% | 1 |
| Duration:≥ 3 months | 4 | 0.02 | -0.43, 0.46 | 0.95 | 0% | 0.94 |
| Frequency:≤ 3 times/week | 5 | 0.04 | -0.34, 0.43 | 0.82 | 0% | 1 |
| Frequency:> 3 times/week | 3 | 0 | -0.45, 0.45 | 0.99 | 0% | 0.86 |
| HIIT interval: 1-3mins | 6 | 0.01 | -0.35, 0.37 | 0.97 | 0% | 1 |
| HIIT interval: ≥3mins | 2 | 0.06 | -0.44, 0.56 | 0.82 | 0% | 0.92 |
| FM |  |  |  |  |  |  |
| Duration:< 3 months | 3 | 0.03 | -0.41, 0.47 | 0.9 | 0% | 0.98 |
| Duration:≥ 3 months | 2 | 0.05 | -0.61, 0.72 | 0.88 | 0% | 0.83 |
| Frequency:≤ 3 times/week | 3 | 0.06 | -0.49, 0.61 | 0.82 | 0% | 0.98 |
| Frequency:> 3 times/week | 2 | 0.02 | -0.47, 0.5 | 0.95 | 0% | 0.9 |
| HIIT interval: 1-3mins | 3 | 0.06 | -0.49, 0.61 | 0.82 | 0% | 0.98 |
| HIIT interval: ≥3mins | 2 | 0.02 | -0.47, 0.5 | 0.95 | 0% | 0.9 |
| FFM |  |  |  |  |  |  |
| Duration:< 3 months | 5 | -0.03 | -0.37, 0.32 | 0.87 | 0% | 0.99 |
| Duration:≥ 3 months | 2 | -0.02 | -0.69, 0.64 | 0.95 | 0% | 0.78 |
| Frequency:≤ 3 times/week | 4 | -0.05 | -0.51, 0.41 | 0.84 | 0% | 0.97 |
| Frequency:> 3 times/week | 3 | -0.01 | -0.42, 0.4 | 0.96 | 0% | 0.98 |
| HIIT interval: 1-3mins | 4 | -0.05 | -0.51, 0.41 | 0.84 | 0% | 0.97 |
| HIIT interval: ≥3mins | 3 | -0.01 | -0.42, 0.4 | 0.96 | 0% | 0.98 |
| HbA1c |  |  |  |  |  |  |
| Duration:< 3 months | 4 | -0.15 | -0.55, 0.24 | 0.45 | 0% | 0.99 |
| Duration:≥ 3 months | 8 | -0.29 | -0.58, 0 | **0.05** | 0% | 0.91 |
| Frequency:≤ 3 times/week | 9 | -0.2 | -0.47, 0.08 | 0.16 | 0% | 1 |
| Frequency:> 3 times/week | 3 | -0.38 | -0.84, 0.09 | 0.11 | 0% | 0.48 |
| HIIT interval: 1-3mins | 7 | -0.29 | -0.62, 0.03 | 0.08 | 0% | 0.86 |
| HIIT interval: ≥3mins | 5 | -0.19 | -0.53, 0.15 | 0.27 | 0% | 0.98 |
| FPG |  |  |  |  |  |  |
| Duration:< 3 months | 4 | 0.16 | -0.22, 0.54 | 0.41 | 0% | 0.67 |
| Duration:≥ 3 months | 8 | -0.01 | -0.3, 0.28 | 0.95 | 0% | 0.92 |
| Frequency:≤ 3 times/week | 7 | -0.04 | -0.35, 0.27 | 0.8 | 0% | 0.97 |
| Frequency:> 3 times/week | 5 | 0.17 | -0.18, 0.51 | 0.34 | 0% | 0.63 |
| HIIT interval: 1-3mins | 7 | 0.02 | -0.3, 0.34 | 0.89 | 0% | 0.88 |
| HIIT interval: ≥3mins | 5 | 0.09 | -0.25, 0.42 | 0.61 | 0% | 0.7 |
| FPI |  |  |  |  |  |  |
| Duration:< 3 months | 4 | -0.16 | -0.54, 0.22 | 0.41 | 0 | 0.63 |
| Duration:≥ 3 months | 5 | -0.2 | -0.71, 0.32 | 0.46 | 51% | 0.09 |
| Frequency:≤ 3 times/week | 5 | -0.09 | -0.49, 0.31 | 0.65 | 13% | 0.33 |
| Frequency:> 3 times/week | 4 | -0.28 | -0.75, 0.19 | 0.25 | 38% | 0.18 |
| HIIT interval: 1-3mins | 3 | -0.38 | -0.86, 0.09 | 0.11 | 0% | 0.67 |
| HIIT interval: ≥3mins | 6 | -0.09 | -0.48, 0.31 | 0.67 | 37% | 0.16 |
| VO2peak |  |  |  |  |  |  |
| Duration:< 3 months | 5 | 0.3 | -0.06, 0.66 | 0.1 | 0% | 0.97 |
| Duration:≥ 3 months | 6 | 0.5 | -0.13, 1.13 | 0.12 | 69% | 0.007 |
| Frequency:≤ 3 times/week | 7 | 0.32 | 0, 0.63 | **0.05** | 0% | 0.46 |
| Frequency:> 3 times/week | 4 | 0.62 | -0.17, 1.41 | 0.12 | 72% | 0.01 |
| HIIT interval: 1-3mins | 6 | 0.48 | 0.13, 0.84 | **0.008** | 0% | 0.95 |
| HIIT interval: ≥3mins | 5 | 0.38 | -0.31, 1.07 | 0.28 | 74% | 0.004 |

Bold indicates significant change ( $p < 0.05$ )
BM, body mass; BMI, body mass index; FM, fat mass; PFM, percent fat mass; FFM, fat-free mass; FPG, fasting plasma glucose; FPI, fasting plasma insulin.

### Sensitivity analysis and publication bias

Heterogeneity was detected in BM(I^2^=41%), BMI(I^2^=35%), FPI(I^2^=19%),and VO2peak (I^2^=41%). After performing a sensitivity analysis by removing each one of the eligible studies, we found the heterogeneity is due to the special forms of HIIT in 1 study. The result of funnel plots and Egger’s tests indicated no indication of publication bias.

## Discussion

Diabetes mellitus is a group of diseases due to insufficient insulin secretion or altered islet function, usually manifested as elevated blood glucose, blood lipids, and a significant increase in BM^[28]^. Patients with T2DM also had a significantly increased cardiovascular disease risk, with risk factors including glucose metabolism, blood lipids, BMI, waist circumference, and FM^[29]^. HIIT is increasingly valued as an emerging form of exercise intervention. Several researchers found that HIIT can promote mitochondrial biogenesis and glucose transport by upregulating peroxidisome proliferator activating receptorγcoactivator-1α(PGC-1a), glucose transporter 4 (GLUT4) and oxidative stress pathway gene expression, thereby improving oral glucose tolerance levels in patients with metabolic diseases^[30][31]^. HIIT can also effectively improve endothelium-dependent relaxation as well as vasodilatory function^[32]^.

As the results showed, though, we did not find significant differences between HIIT and MICT on BM, BMI, PFM, FM, FFM, FPG, and FPI in people with T2DM, which was similar to the previous analysis^[33]^. HIIT was found to be superior to MICT in improving HbA1c, and VO_2peak_. As the subgroup analysis indicated, there were statistical differences between HIIT and MICT on HbA1c in long-term(≥3 months) intervention, while in the short-term(<3 months) were not found. Given intervention characteristics, compared to MICT, <3 times/week and 1-3 mins interval HIIT might bring more positive influences on CRF in T2DM respectively. No matter duration, frequency, and HIIT interval, there were no significant differences were found between HIIT and MICT on body composition in T2DM. These findings might be of great help in strategy designing to improve body composition, CRF, and glouse control in T2DM.

Relative to body composition, Improving body composition is closely related to controlling blood glucose and reducing blood lipid and blood pressure in patients with T2DM. Increased BMI can increase the risk of atherogenesis. A study indicated that for T2DM patients with BMI> 25 Kg/m^2^, every 5 Kg/m^2^ increase in BMI means a 40% increase in cardiovascular mortality^[34]^. Although recent studies found that both HIIT and MICT can improve body composition significantly, there were no significant differnces between HIIT and MICT on body composition^[35][36][37]^, which is similar to our results. However, some studies suggest the opposite conclusion: Karstoft^[38]^ et al. found that HIIT is better than MICT in reducing BM, BMI and improving waist-to-hip ratio. The mechanism why HIIT improves body composition in patients with T2DM compared to MICT may be that: First, HIIT promotes energy consumption during or after exercise, and total energy expenditure plays a key role in improving body composition. Scribbans^[39]^et al. proposed that the low-intensity interval training stage interspersed in HIIT belongs to aerobic training, which increases the ratio of fat decomposition, while the high-intensity exercise stage increases the fatigue resistance of muscles, thus prolonging the time of fat decomposition, and HIIT can cause changes in body function and then increase energy consumption and fat decomposition. Second, HIIT inhibits appetite. A recent study shown that HIIT may promote the central inhibition of the brain’s desire to eat high-energy foods through high-intensity exercise, and is able to regulate the hormone levels related to feeding function, which can achieve rapid weight loss in a short period of time^[40]^. Nevertheless, the effects of HIIT and MICT on body composition in patients with T2DM remains controversial. On the one hand, the improvement in body composition in patients with T2DM is influenced by diet and lifestyle. Wu^[41]^ et al. indicated that mere physical activity in the absence of calorie restriction does not lead to meaningful body composition improving. On the other hand, the differences in exercise intensity, exercise frequency, exercise style, and the studied population are the key factors affecting the body composition, although our subgroup analysis did not find duration, frequency, and HIIT interval can make HIIT bring more significant improvement than MICT on body composition in T2DM. For the same intervention period and exercise style, in terms of improving body composition, the time per exercise in the HIIT group was only half the time of a single exercise in the MICT group, which saved patients’ time to some extent. HIIT is also proved to be a well-adapted exercise and HIIT stimulates the secretion of catecholamine lipids is more effective than MICT, the increase of hormone levels strengthened the fat oxidation, and greater fat oxidation may be indirectly associated with sympathetic nervous system activation, can promote the increase of circulating free fatty acid levels after exercise^[42]^. Therefore, HIIT should be used to improve body composition for T2DM people with low sedentary time, overweight / obese and lack of time.

As an independent measure of exercise parameters, CRF is very important for human health. Improving physical activity can significantly enhance cardiopulmonary endurance. A high level of cardiopulmonary endurance can effectively reduce the risk of T2DM cardiovascular disease and improve the quality of life^[43]^. Our results found that HIIT can bring more improvement on VO_2peak_ than HIIT in T2DM. Similar results were found in a recent study, both the HIIT and MICT could increase VO_2peak_, and the HIIT was better to the MICT in improving cardiopulmonary endurance indicators^35^. Støa^[44]^ et al found that after 12 weeks intervention a significant increase in HIIT VO_2peak_ and VO_2peak_ /kg, with no significant change in MICT group before and after intervention, and no significant difference in improving lactate threshold. However, a recent study found that there is no difference between the HIIT and MICT groups in the VO2peak group with the same total control load^[45]^. CRF is the main factor determining the strength of exercise capacity in T2DM, and although the current study cannot determine the physiological mechanism of HIIT improving CRF, it is speculated that the effects of HIIT may involve central and peripheral adaptations, including increased cardiac output, improved vascular / endothelial function, and increased muscle oxidation^[46][47]^.

HbA1c is less affected by drugs and has high stability, so HbA1c can be used as a diagnostic index in the evaluation of blood glucose index in patients with T2DM. HbA1c Researchers proved that for each 11% decrease, the complication rate of T2DM decreased by 25%^35^. Although our reuslts showed that HIIT is superiar than MICT in improving HbA1c, the effects of HIIT on glucose control in T2DM are debated. Winding^35^ et al found that FPG and HbA1c decreased significantly after intervention, but the difference was not statistically significant compared with the MICT group, which showed similar results in this study, which is consistent with our results. This may be because the intervention programs are different. In a recent study, patients took 85% -90% HRpeak in 1min and 1 min low intensity exercise for 4 weeks, and was found that HIIT was better than the MICT group in reducing blood glucose^[48]^. Umpierre^[49]^ et al found that HIIT improves blood glucose control more than low intensity aerobic exercise, when controlling energy expenditure. There were no significant diffiences found in our results between HIIT and MICT on FPG and FPI in T2DM. However, HIIT was proved to be more strongly increased glucose transporter GLUT-4 expression in skeletal muscle, and transporter GLUT-4 expression accelerated the process of glucose metabolism^9^. Whether HIIT has more benefits on glucose control than other aerobic exercise in T2DM still need more evidence.

Although we conducted a comprehensive study on the effect of HIIT and MICT on body compositon and glucose control in people with T2DM through meta-analysis and subgroup-analysis, this review still has several limitations. As a result of strict inclusion and exclusion criteria, the subjects of all the eligible studies were limited(<20). And it was unclear whether HIIT is finished by subjects without supervision. A recent study^[50]^ indicated that supervision of exercise can positively improve the effects of HIIT, while our study did not include it as a covariable. In addition, the dietary control of participants was misanalyzed in this study which might influence the outcomes, as several studies have proven that the combination of exercise and diet intervention brings more effects on body composition^[51],[52]^. Finally, notwithstanding statistical differences found in this meta-analysis, relative to MICT, the improvement HIIT brings was so limited that whether HIIT has more clinical meanings on HbA1c and VO_2peak_ is hard to say. Future studies must pay more attention to the forms of HIIT and the combination with dietary intervention to expand the clinical significance of HIIT in T2DM. Studies with more sample size, longer intervention, and better assessment need to be conducted to provide more compelling evidence to elucidate the time-saving and efficiency of HIIT.

## Conclusions

HIIT provides more benefits on VO_2peak_ and HbA1c relative to MICT, which might be influenced by many factors, including duration(≥3 months), frequency, and HIIT interval. For people with T2DM, HIIT can achieve more improvement in CRF and glucose control than MICT. In summary, compared to MICT, our study indicated that the advantages HIIT brings to T2DM on body composition and glucose control are limited, yet these benefits can be provoked in a more time-saving manner.

## Data Availability

All relevant data are within the manuscript and its Supporting Information files.

---

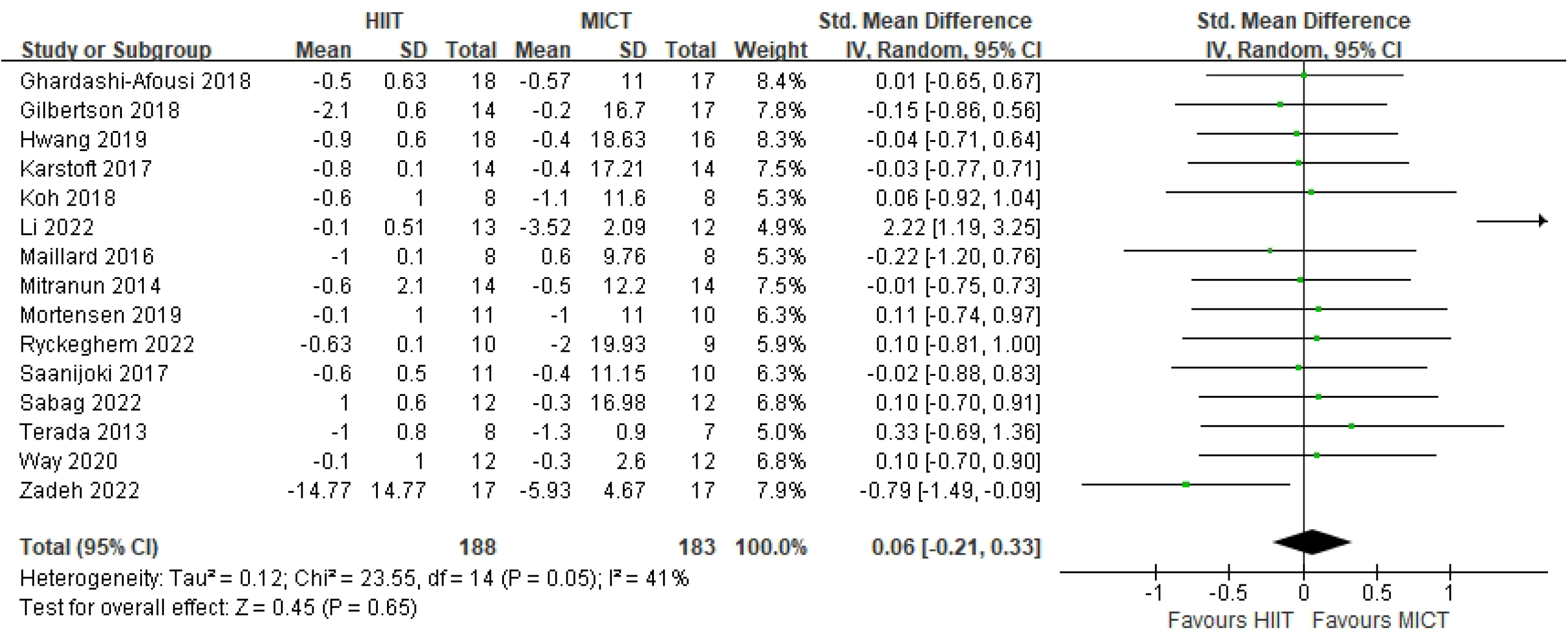

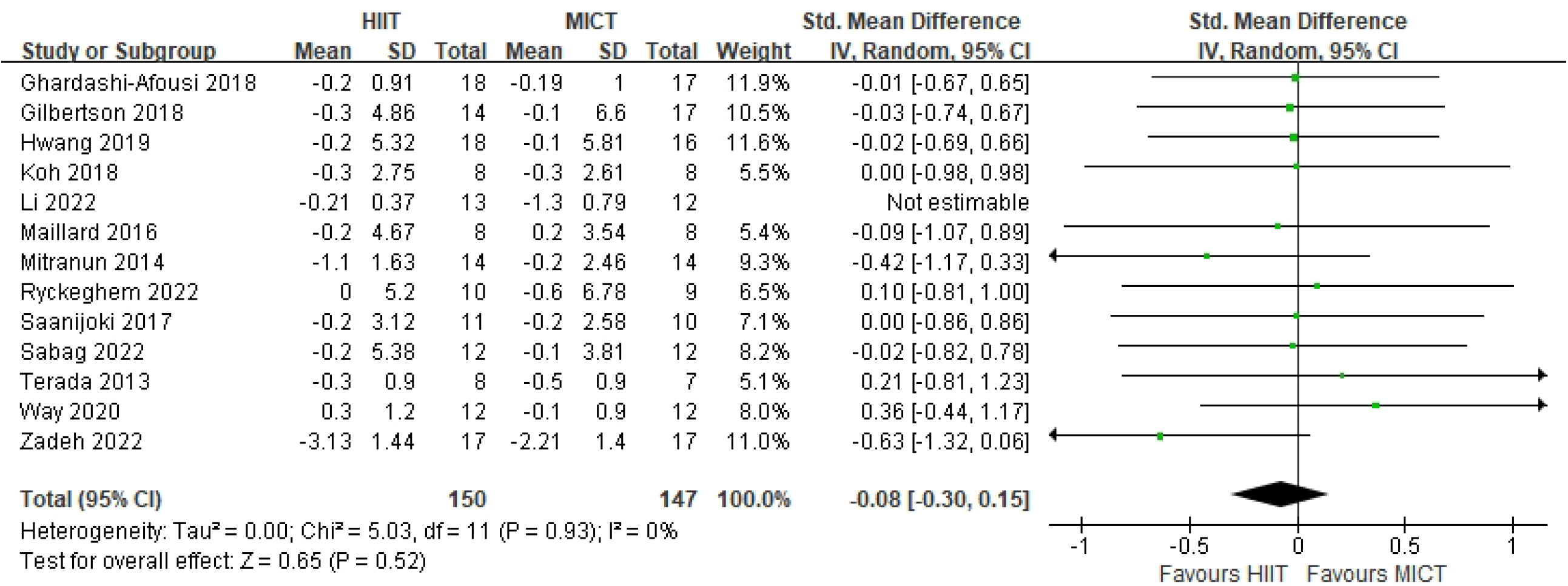

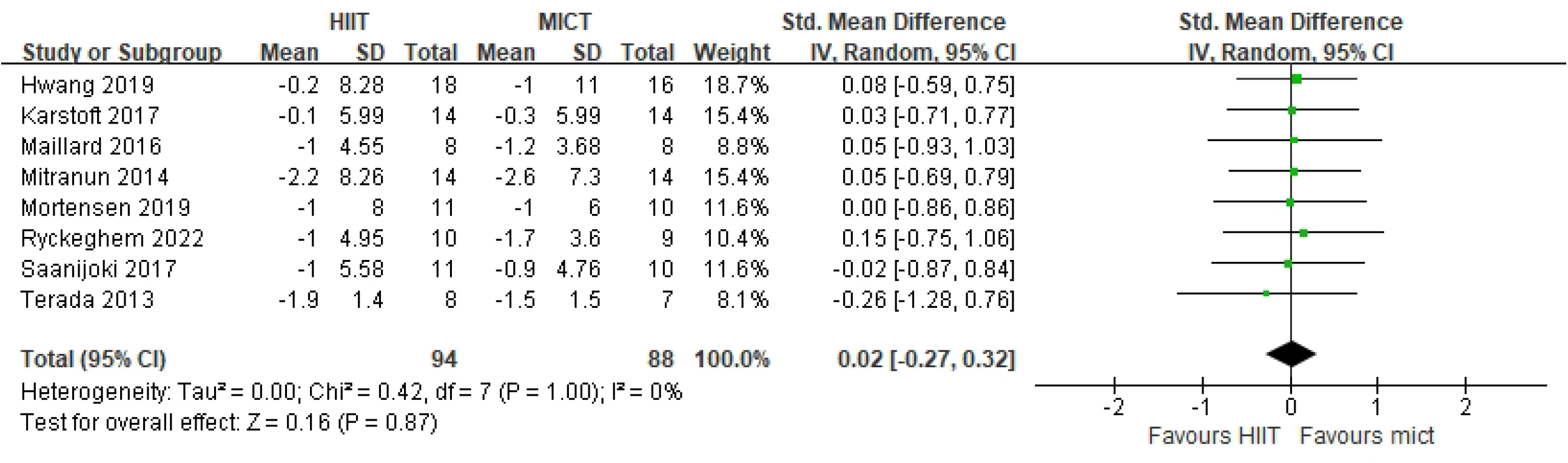

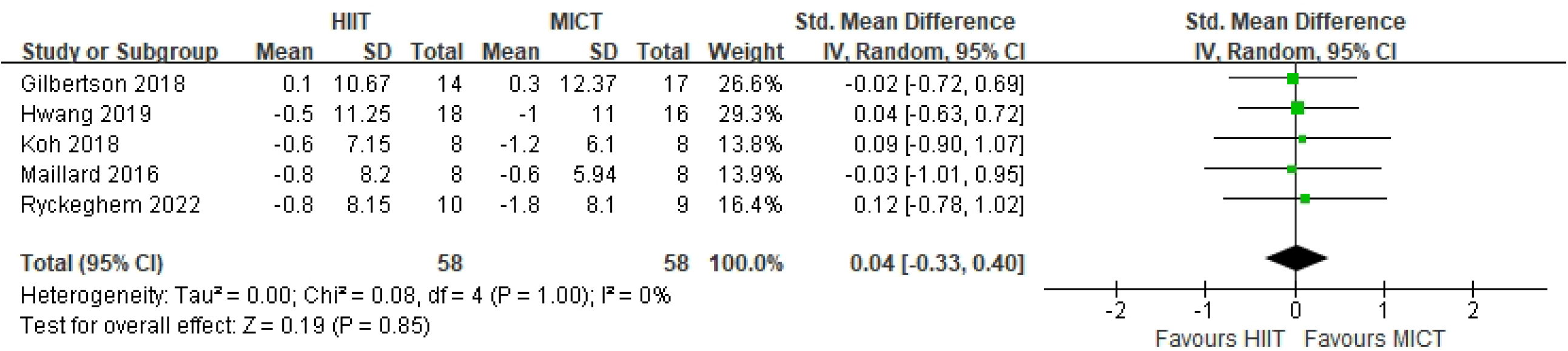

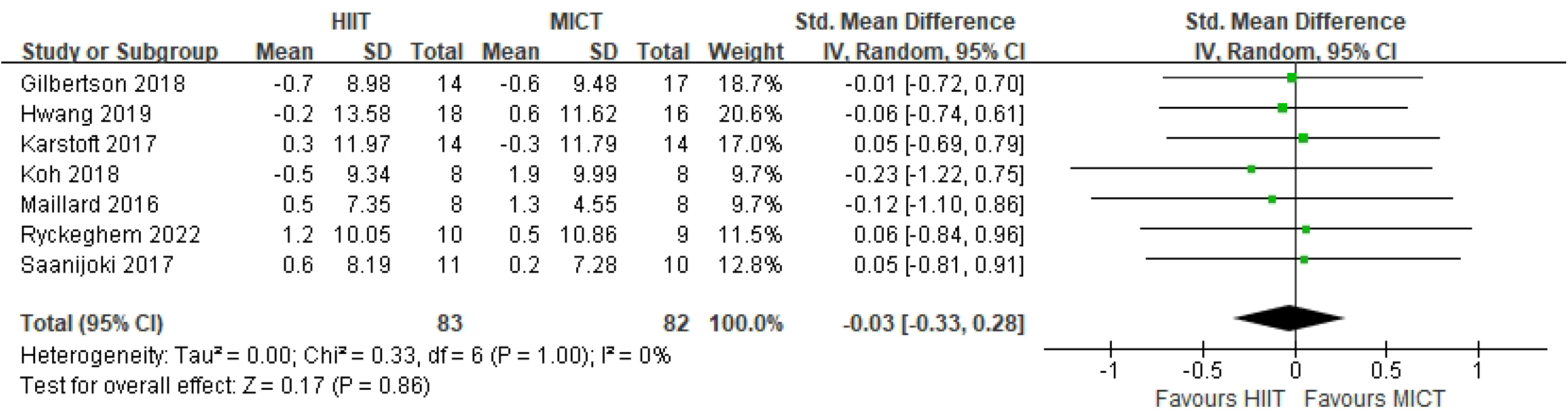

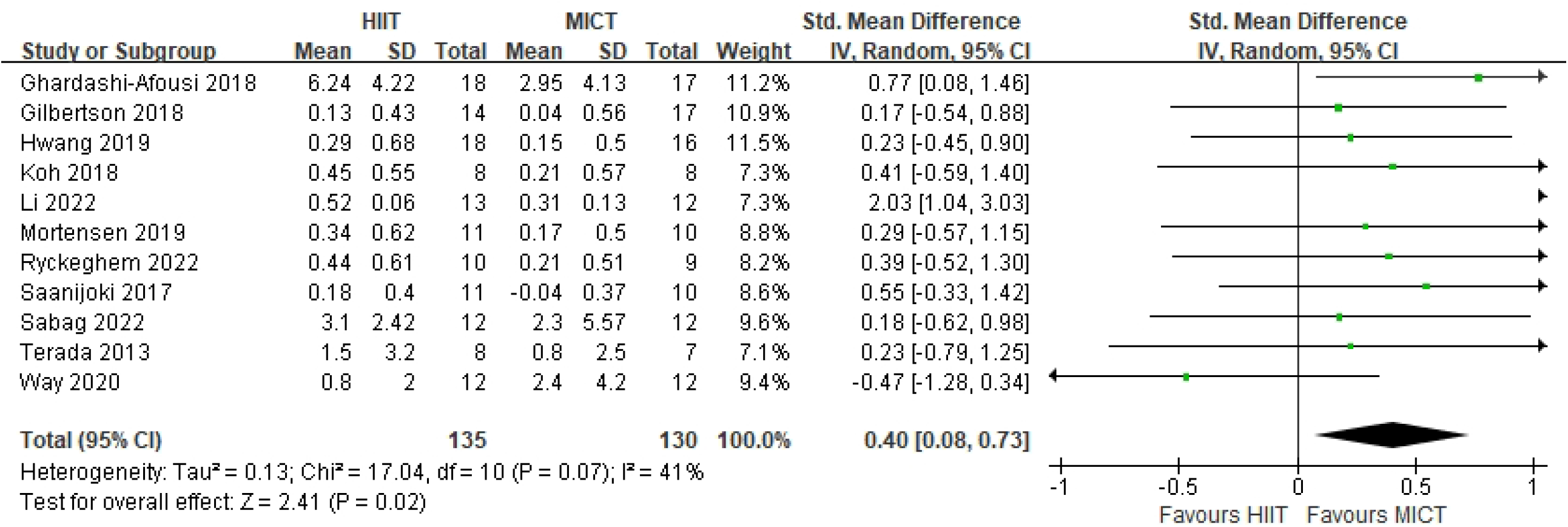

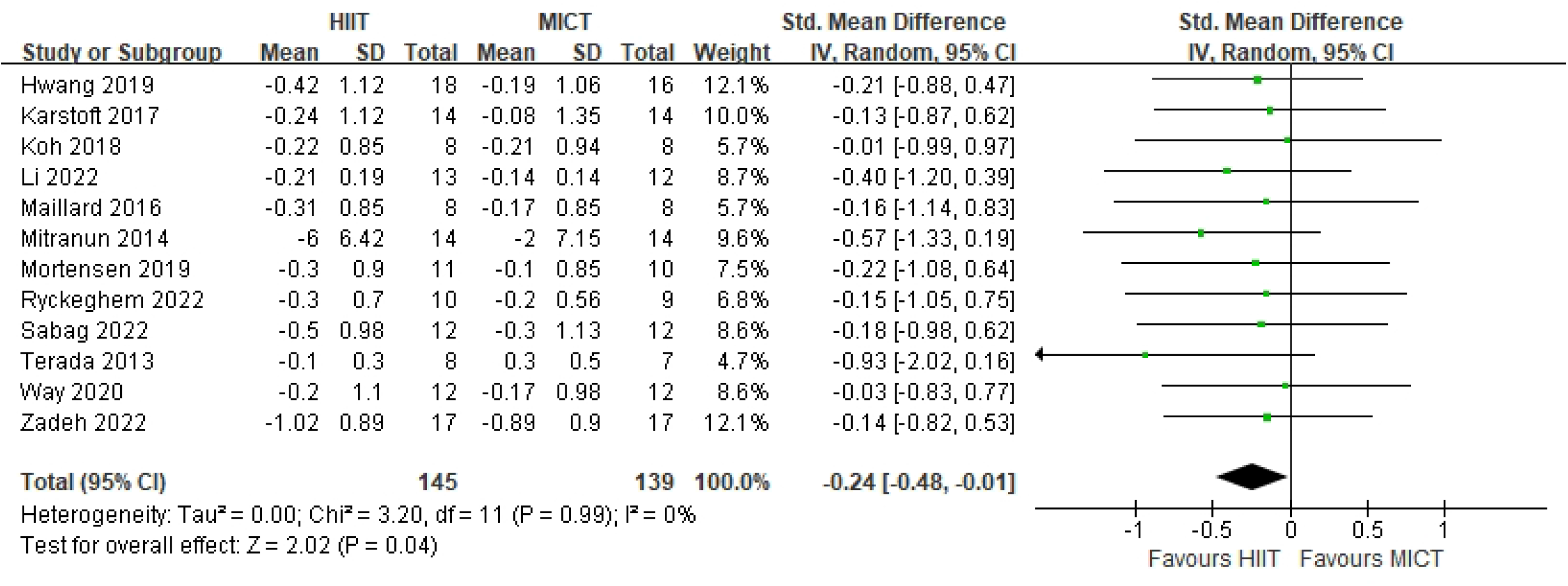

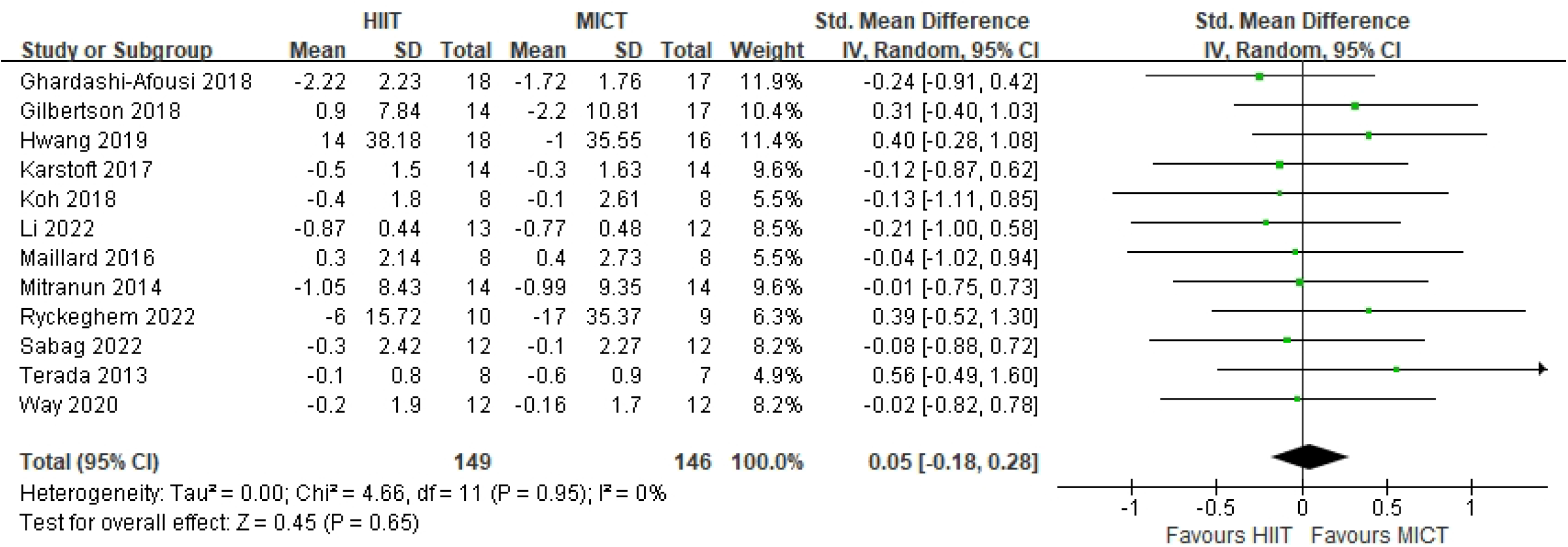

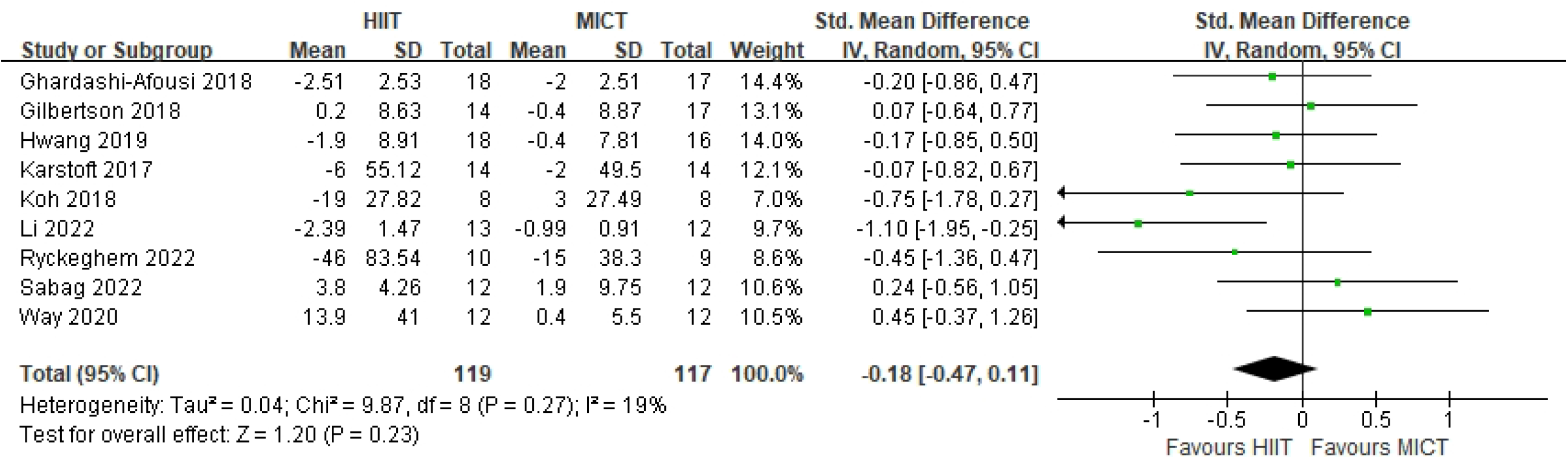

